# Association of cannabis use-related predictor variables and self-reported psychotic disorders: US adults, 2001-2002 and 2012-2013

**DOI:** 10.1101/2020.09.25.20201871

**Authors:** Ofir Livne, Dvora Shmulewitz, Aaron L. Sarvet, Deborah S. Hasin

## Abstract

**Objective:** To determine the association of cannabis use-related variables and self-reported psychotic disorders during two time periods (2001-2002; 2012-2013).

**Methods:** Logistic regression was used to analyze data from the National Epidemiologic Survey on Alcohol and Related Conditions (NESARC, 2001-2002; N=43,093) and NESARC-III (2012-2013; N=36,309). Among those with and without cannabis predictors (any and frequent [≥3 times a week] non-medical use, DSM-IV cannabis use disorders [CUD], cannabis dependence [CD]), standardized prevalence of past-year self-reported psychotic disorders were estimated. Association was indicated by within-survey differences in psychotic disorders by cannabis-related predictor status. Whether associations changed over time was indicated by difference-in-difference tests (contrasts between the surveys).

**Results:** In both surveys, self-reported psychotic disorders were significantly more prevalent in those with than those without any non-medical cannabis use (2001-2002: 1.65% vs 0.27%; 2012-2013: 1.89% vs. 0.68%), with similar associations in both periods. Self-reported psychotic disorders were unrelated to frequent non-medical use in 2001-2002 but were significantly more prevalent in those with than without frequent non-medical use in 2012-2013 (2.68% vs. 0.71%), with no significant difference over time. In both surveys, self-reported psychotic disorders were significantly more prevalent in those with than without CUD (2001-2002: 2.43% vs. 0.30%; 2012-2013: 3.26% vs. 0.72%), with no significant differences in the associations over time. Self-reported psychotic disorders were unrelated to CD in 2001-2002 but were significantly more prevalent in those with than without CD in 2012-2013 (8.54% vs. 0.73%), showing a significantly stronger relationship in 2012-2013; similarly, among past-year non-medical cannabis users, the association was significantly stronger in 2012-2013.

**Conclusions:** Cannabis-related variables, especially cannabis dependence, remain related to self-reported psychotic disorders. Therefore, clinicians should closely monitor cannabis-dependent users and assess the need for preventive and therapeutic interventions for these individuals.

## INTRODUCTION

Schizophrenia spectrum and other psychotic disorders are a heterogenous group of serious mental health disorders that involve impairment in thinking, perception and emotion(1, 2). Despite being relatively uncommon in the general population, psychotic disorders result in substantial social, economic, and health-related burdens(1, 3), are leading causes of disability-adjusted life-years in the US and worldwide(4-6), and increase the risk of suicide and early mortality(7-9). Past studies that examined whether the incidence and prevalence of psychotic disorders have changed over time have produced mixed results with marked heterogeneity in time trends between different types of psychotic disorders(10). Most of the data utilized in these studies originated in samples outside of the US, mainly from northern Europe and the UK; findings from these studies indicated a decline or no change in incidence or prevalence rates of psychotic disorders over time(10-17). Nevertheless, most of these studies are older, from nearly 15-20 years ago. In the US, data from the US Healthcare Cost and Utilization Project(18), which focuses on inpatient discharge diagnoses of psychosis, indicate that rates of psychotic disorders have increased by over 40% from 2004 to 2016 in the US. Further, using data from the US national Veterans Affairs system, one study showed that the prevalence of patients treated for psychotic disorders increased between 2007 and 2013(19). Further studies utilizing large-scale, national data are needed to further understand changes in rates of psychotic disorders in the US general population.

The etiology of psychotic disorders has been widely studied over the years and shown to be multifactorial, involving heritable and socioenvironmental factors (e.g., childhood adversities, social disadvantages, and poor social networks)(20, 21). While the relationship between cannabis use and psychosis has been debated in recent decades, reviews and meta-analyses(22-24) indicate that use of cannabis, the most widely used illicit substance in the US and worldwide(25), is a causal factor in the risk of developing psychosis. Furthermore, there is evidence of a dose-response relationship of frequency of cannabis use(26, 27) and potency of cannabis(28) with incidence of psychotic disorders. Continued use of cannabis has also been linked to longer hospitalizations and frequent relapses among individuals with psychotic disorders(29, 30). Studies focusing on associations between cannabis dependence (CD) and psychosis are less frequent than those examining cannabis use; however, findings indicate that cannabis dependence is a risk factor for development of psychotic disorders(31) and that a diagnosis of cannabis dependence at an early age is associated with an early age at onset of psychosis(32). Nevertheless, only one(33) of the studies reporting the abovementioned associations used data that were representative of the US general population, nor did they include in their analyses the more recent DSM-5 diagnostic criteria for cannabis use disorder (CUD).

Overall, the prevalence of adult cannabis use and of CUD has increased across several US national studies in recent years(34, 35). A growing number of studies show that medical cannabis laws (MCL) and recreational cannabis laws (RCL) have a causal role in these observed increases(36-38). As additional states consider MCL and RCL, more attention should be put on the potential health consequences, including an increased risk for psychotic disorders. To-date, studies investigating changes over time in the association between cannabis use, CUD and psychotic disorders in the US population are lacking.

Using data from two US nationally representative adult surveys, the 2001-2002 National Epidemiologic Survey on Alcohol and Related Conditions (NESARC) and the 2012-2013 NESARC-III, we examined whether: 1) prevalence of psychotic disorders changed over time (i.e., between the two surveys); 2) any or frequent non-medical cannabis use, CD, or CUD were associated with the prevalence of psychotic disorders in either survey; and 3) these relationships changed from 2001-2002 to 2012-2013, either overall or in a subgroup including only past-year cannabis users.

## METHODS

### Samples and procedures

The NESARC (39) and NESARC-III surveys (40) used multistage designs to sample adults (age ≥18) in households and group quarters. Sample weights adjusted for nonresponse and probability of selection. The total sample analyzed was 79,402 (43,093 in NESARC, 36,309 in NESARC-III). Across surveys, rigorous field procedures were similar (35, 37), including structured in-class training and home-study for interviewers, random callbacks to verify interview data, and expert supervision. The examination of trends over time in important health outcomes was possible due to the methodological similarity between the two surveys (35, 41-43). For the 2001-2002 NESARC, the US Bureau of the Census and Office of Management and Budget institutional review boards (IRB) approved the protocol and written consent procedures. Response rate for NESARC (2001-2002) was 81.0%. For the NESARC-III, the IRB at the National Institutes of Health and Westat approved the protocol and verbal (recorded electronically) consent procedures. Response rate for NESARC-III was 60.1%, similar to other US representative surveys conducted in similar years (44, 45).

### Measures

In both surveys, substance use and substance use disorders were assessed using a structured computer-assisted interview, the Alcohol Use Disorder and Associated Disabilities Interview Schedule (AUDADIS).

The outcome, self-reported psychotic disorders over the past year, was measured in NESARC and NESARC-III with a nearly identical question, asking if a doctor or other health professional informed the respondent that they had schizophrenia or psychotic illness or episode. Prevalences of schizophrenia in North America reported in studies that used self-reported measurements are consistent with those reported in structured interview designed studies(46).

Predictors included four past-year cannabis use-related variables: any non-medical use, frequent non-medica use, DSM-IV cannabis dependence (CD), and DSM-IV CUD In both surveys, identical questions were used to assess non-medical cannabis use, defined as use without a prescription or other than prescribed, e.g., to get high (37). Any use was positive for those who used ≥1 time in the past year; frequent use was positive for those who used ≥3 times per week. Dependence was positive for those endorsing ≥3 of 6 DSM-IV dependence criteria (cannabis withdrawal was not included in DSM-IV). CUD was positive for those with DSM-IV cannabis dependence or abuse, which was positive for those endorsing ≥1 of 4 DSM-IV abuse criteria. Abuse and dependence were combined because their criteria reflect a single disorder (47). In both surveys, the 22 CUD symptom items were mostly identical; the large differences in CUD prevalence across the two surveys could not be accounted for by the few slight differences in item wording (35, 37). For sensitivity analyses, we re-defined CD and CUD, adding a cannabis withdrawal criterion and requiring 3 of 7 dependence criteria to be positive for CD. Cannabis withdrawal was assessed identically in NESARC and NESARC-III, as 3 or more of 5 withdrawal symptoms: nervousness/anxiety, sleep difficulty, depressed mood, restlessness, or physical symptoms (one or more of headache, shakiness, sweating, abdominal pain, and fever); or use to avoid or relieve withdrawal symptoms. This was done because DSM-5 includes withdrawal as a CUD criterion, due to evidence showing its validity and relatedness to the other CUD criteria (47).

Control covariates included gender; age (18-29; 30-44; 45-64; ≥ 65 years); race/ethnicity (Hispanic; Non-Hispanic: White; Black; and Other [Native American, Asian, Pacific Islander]); education (<high school; high school graduate or GED; ≥ some college); and urbanicity (urban, rural). In sensitivity analyses, we included a covariate indicating if respondents’ states of residence had a MCL, as determined by economic and legal experts as in previous papers (37, 48, 49). Seven states (California, Colorado, Hawaii, Maine, Nevada, Oregon, Washington) had MCL by 2001 (NESARC). These and nine more states (Arizona, Connecticut, Maryland, Massachusetts, Michigan, Montana, New Jersey, New Mexico, Vermont) had MCL by 2012 (NESARC-III).

### Statistical Analysis

As in other studies evaluating trends between the two surveys (35, 37, 41, 42), the NESARC and NESARC-III datasets were concatenated, adding a survey variable. To determine the change in self-reported psychotic disorders over time, logistic regression was used to model self-reported psychotic disorders as a function of survey (time) and sociodemographic control variables (age, race/ethnicity, gender, education, and urbanicity). Model-predicted standardized prevalence (i.e., back-transformed from the log scale with sociodemographics averaged between the surveys) of self-reported psychotic disorders was estimated in each survey; the difference between the prevalence estimates indicated the change over time.

Logistic regression was then used to evaluate the association of each cannabis-related predictor variable and self-reported psychotic disorders within each survey and whether those associations changed between the surveys, modeling self-reported psychotic disorders as a function of each cannabis-related predictor, survey, cannabis-related predictor x survey interactions, and sociodemographic control variables. Model-predicted standardized prevalence of self-reported psychotic disorders was estimated in each survey by cannabis-related predictor status (yes vs. no). The difference in these prevalence estimates indicated association of the cannabis-related predictor with self-reported psychotic disorders, within each survey. Whether the associations differed between the surveys (i.e., changed over time) was indicated by contrasts between these prevalence differences (difference-in-difference tests). The association models were analyzed in the whole sample and among past-year non-medical cannabis users (N=5,304; 1,603 from NESARC and 3,701 from NESARC-III). Additive effects and interactions were evaluated because those are considered most appropriate for assessing the public health significance of interactions (50-52), since the measures of differences are proportional to the number of excess cases in the exposed group (those with the cannabis-related predictor). For readers more familiar with relative risks, we also evaluated effects and interactions on the multiplicative scale. Using the logistic regression models described above, relative risk was indicated by the prevalence ratio, i.e. the prevalence of self-reported psychotic disorders in those with the cannabis-predictor divided by prevalence in those without, within each survey. Multiplicative interaction was evaluated as the ratio of prevalence ratios, i.e., prevalence ratio for 2012-2013 divided by the prevalence ratio for 2001-2002.

For all analysis, SUDAAN 11.0.1 (53) was used, incorporating survey weights to adjust for the complex sampling design, to yield US adult population-representative estimates. Statistical tests were 2-tailed with significance based on p<0.05, as indicated by 95% confidence intervals (CI). Interpretation of the 95% CI differs for difference (additive) and relative (multiplicative) effects. For differences, a value of 0.0 indicates no difference, so an estimate with 95% CI not including 0.0 is statistically significant at p<0.05. For ratios, a value of 1.0 indicates no difference, so an estimate whose 95% CI do not include 1.0 is statistically significant at p<0.05.

Several sensitivity analyses were conducted. First, to reflect the addition of cannabis withdrawal to the DSM-5, we added cannabis withdrawal to the dependence criteria and re-ran the models for CD and CUD. Second, we added a covariate indicating state MCL status at the time of each survey and re-ran the models. Participants from the 42 states included in both surveys were included in this analysis (41,706 from NESARC; 36,309 from NESARC-III; total=78,015), as in previous studies (37). Third, we ran a model, adding an MCL variable in 3-way interactions with each cannabis-related predictor and survey, to explore whether associations between the cannabis-related variables and self-reported psychotic disorders were more likely to change over time among those in MCL states. The MCL variable was defined with three levels: never-MCL; MCL enacted by 2001; and MCL enacted between 2002 and 2012.

## RESULTS

### Trend in self-reported psychotic disorders

The standardized prevalence of past-year self-reported psychotic disorders among US adults increased significantly from 0.33% in 2001-2002 to 0.80% in 2012-2013, for an increase of +0.47% (95% CI=0.33, 0.61), and a prevalence ratio of 2.45 (95% CI=1.88, 3.19).

### Association with self-reported psychotic disorders

#### Any past-year non-medical cannabis use

The standardized prevalence of self-reported psychotic disorders was significantly greater in those with any non-medical cannabis use vs those without use in both periods (Table 1: 2001-2002: 1.65% vs 0.27%; 2012-2013: 1.89% vs. 0.68%). The difference-in-difference test indicated that the association strength did not differ significantly between 2012-2013 and 2001-2002.

**Table 1.** Association of past-year cannabis use-related variables with self-reported psychotic disorders, by survey

| Predictors: past-year cannabis use-related variables | Whole sample |  | Among past-year cannabis users |  |
| --- | --- | --- | --- | --- |
|  | 2001-2002<br>(NESARC)<br>N=43,093 | 2012-2013<br>(NESARC-III)<br>N=36,309 | 2001-2002<br>(NESARC)<br>N=1,603 | 2012-2013<br>(NESARC-III)<br>N=3,701 |
| **Any non-medical cannabis use<sup>a</sup>** |  |  |  |  |
| With any cannabis use (prevalence<sup>b</sup> [SE]) | 1.65 (0.47) | 1.89 (0.33) |  |  |
| Without any cannabis use (prevalence<sup>b</sup> [SE]) | 0.27 (0.03) | 0.68 (0.06) |  |  |
| **Prevalence difference<sup>c</sup> (95% CI)** | *1.38 (0.47, 2.29)* | *1.21 (0.56, 1.86)* |  |  |
| **Difference in prevalence differences<sup>c</sup> (95% CI)** | reference | -0.17 (-1.24, 0.90) |  |  |
| Prevalence ratio<sup>d</sup> (95% CI) | *6.04 (3.42, 10.66)* | *2.78 (1.90, 4.06)* |  |  |
| Ratio of prevalence ratios<sup>d</sup> (95% CI)<sup>f</sup> | reference | 0.46 (0.24, 1.10) |  |  |
| **Frequent non-medical cannabis use** |  |  |  |  |
| With frequent cannabis use (prevalence<sup>b</sup> [SE]) | 0.94 (0.43) | 2.68 (0.60) | 2.01 (0.75) | 3.78 (0.75) |
| Without frequent cannabis use (prevalence<sup>b</sup> [SE]) | 0.32 (0.03) | 0.71 (0.06) | 2.85 (0.81) | 2.09 (0.38) |
| **Prevalence difference<sup>c</sup> (95% CI)** | *0.62 (-0.21, 1.45)* | *1.96 (0.79, 3.13)* | *-0.84 (-2.98, 1.31)* | *1.68 (-0.05, 3.41)* |
| **Difference in prevalence differences<sup>c</sup> (95% CI)** | reference | 1.34 (-0.06, 2.74) | reference | 2.52 (-0.17, 5.21) |
| Prevalence ratio<sup>d</sup> (95% CI) | *2.96 (1.25, 6.99)* | *3.75 (2.37, 5.93)* | *0.71 (0.28, 1.77)* | *1.80 (1.03, 3.17)* |
| Ratio of prevalence ratios<sup>d</sup> (95% CI)<sup>f</sup> | reference | 1.27 (0.53, 8.84) | reference | 2.54 (0.89, 28.98) |
| **DSM-IV Cannabis Use Disorder (CUD)<sup>e</sup>** |  |  |  |  |
| With DSM-IV CUD (prevalence<sup>b</sup> [SE]) | 2.43 (1.03) | 3.26 (0.83) | 3.65 (1.43) | 4.36 (0.93) |
| Without DSM-IV CUD (prevalence<sup>b</sup> [SE]) | 0.30 (0.03) | 0.72 (0.06) | 2.09 (0.58) | 2.09 (0.37) |
| **Prevalence difference<sup>c</sup> (95% CI)** | *2.13 (0.12, 4.15)* | *2.54 (0.92, 4.17)* | *1.56 (-1.38, 4.51)* | *2.26 (0.18, 4.35)* |
| **Difference in prevalence differences<sup>c</sup> (95% CI)** | reference | 0.41 (-2.06, 2.87) | reference | 0.70 (-2.83, 4.22) |
| Prevalence ratio<sup>d</sup> (95% CI) | *8.20 (3.59, 18.70)* | *4.53 (2.69, 7.63)* | *1.75 (0.70, 4.36)* | *2.08 (1.15, 3.78)* |
| Ratio of prevalence ratios<sup>d</sup> (95% CI)<sup>f</sup> | reference | 0.55 (0.22, 3.12) | reference | 1.19 (0.40, 13.06) |
| **DSM-IV Cannabis Dependence (CD)** |  |  |  |  |
| With DSM-IV CD (prevalence<sup>b</sup> [SE]) | 2.36 (1.27) | 8.54 (2.36) | 3.95 (1.93) | 10.91 (2.90) |
| Without DSM-IV CD (prevalence<sup>b</sup> [SE]) | 0.32 (0.03) | 0.73 (0.06) | 2.52 (0.67) | 2.08 (0.30) |
| **Prevalence difference<sup>c</sup> (95% CI)** | *2.04 (-0.45, 4.53)* | *7.81 (3.19, 12.42)* | *1.43 (-2.54, 5.40)* | *8.83 (3.06, 14.60)* |
| **Difference in prevalence differences<sup>c</sup> (95% CI)** | reference | *5.77 (0.67, 10.87)* | reference | *7.40 (0.46, 14.34)* |
| Prevalence ratio<sup>d</sup> (95% CI) | *7.39 (2.57, 21.24)* | *11.70 (6.67, 20.51)* | *1.57 (0.53, 4.64)* | *5.24 (2.82, 9.75)* |
| Ratio of prevalence ratios<sup>d</sup> (95% CI)<sup>f</sup> | reference | 1.58 ** | reference | 3.34 ** |

a Non-medical cannabis use: without a prescription or other than prescribed, e.g., to get high.
b Predicted prevalence of self-reported psychotic disorders, adjusted for sociodemographic covariates (age, gender, race/ethnicity, education level, and urbanicity), from logistic regression
c Effects estimated on the additive scale: **prevalence difference** indicates the prevalence difference of psychotic disorders between those with and without the cannabis predictor in 2001-2002 and 2012-2103, while **difference in prevalence differences** indicates the difference between those differences. These effects are significant when the 95% confidence interval (CI) does not include 0.
d Effects estimated on the multiplicative scale: **prevalence ratio** indicates the ratio of the prevalence of psychotic disorders between those with and without the cannabis predictor in 2001-2002 and 2012-2103, while **ratio of prevalence ratios** indicates the ratio between those ratios. These effects are significant when the 95% CI does not include 1.
e DSM-IV dependence or abuse
<sup>f</sup> calculated using the Fieller Method (https://www.graphpad.com/quickcalcs/errorProp1/?Format=SEM); \*\* 95% CI could not be calculated
Significant effects are italicized

#### Frequent non-medical cannabis use

Frequent non-medical cannabis use was not associated with self-reported psychotic disorders in 2001-2002 (Table 1). Although the standardized prevalence of self-reported psychotic disorders was significantly greater in those with vs. without frequent non-medical use by 2012-2013 (2.68% vs. 0.71%), the difference-in-difference test did not indicate a significantly stronger association in 2012-2013 than in 2001-2002. Among past-year non-medical cannabis users, the association in 2012-2013 became non-significant (Table 1).

#### DSM-IV cannabis use disorder (CUD)

The standardized prevalence of self-reported psychotic disorders was significantly greater among those with DSM-IV CUD vs those without in both periods (Table 1: 2001-2002: 2.43% vs. 0.30%; 2012-2013: 3.26% vs. 0.72%). The difference-in-difference test indicated that the association strength did not differ significantly between 2012-2013 and 2001-2002. Among past-year users (Table 1), significant association was observed in 2012-2013 but not in 2001-2002, with no observed difference between association strength.

#### DSM-IV cannabis dependence (CD)

CD was not associated with self-reported psychotic disorders in 2001-2002 (Table 1). By 2012-2013, the standardized prevalence of self-reported psychotic disorders was significantly greater in those with vs. without CD (8.54% vs. 0.73%). The difference-in-difference test indicated significantly stronger association of self-reported psychotic disorders and CD in 2012-2013 than in 2001-2002. Similar results were observed among past-year users (Table 1), with a significantly stronger association in 2012-2013 than in 2001-2002.

### Sensitivity analyses

After adding cannabis withdrawal to the dependence criteria, both CUD and CD were associated with self-reported psychotic disorders in both time periods, with no differences in the association strength between 2012-2013 and 2001-2002; similar results were observed with cannabis withdrawal (Supplemental Table 1). Among past-year non-medical cannabis users, CUD and CD were significantly associated with self-reported psychotic disorders in 2012-2013, but the associations did not differ significantly over time (Supplemental Table 1).

When adding MCL as a covariate, results were virtually the same as those from the original models, overall and among past-year non-medical cannabis users (Supplemental Table 2), except that frequent use was associated with self-reported psychotic disorders among users in 2012-2013. No test exploring change in the associations of the cannabis-related predictors and self-reported psychotic disorders over time by state MCL status was significant (Supplemental Table 3).

### Relative scale

On the relative scale, any past year non-medical cannabis use, frequent use, DSM-IV CUD, and CD were all significantly associated with self-reported psychotic disorders in both 2001-2002 and 2012-2013, with no change in magnitude of association over time (Table 1). Among past-year non-medical cannabis users, frequent use, CUD, and CD were significantly associated with self-reported psychotic disorders in 2012-2013, with no change in magnitude of association over time (Table 1). When withdrawal was added, both CUD and CD were significantly associated with self-reported psychotic disorders in both time periods, with no significant differences in the association strength, overall and among users (Supplemental Table 1). Results after adding MCL as a covariate were the same as the main results (Supplemental Table 2).

## DISCUSSION

The current study examined associations between several cannabis use-related variables and self-reported psychotic disorders, and changes over time in these associations, in the general adult US population. In recent decades there have been substantial shifts in the US cannabis landscape, including increased perception of cannabis as a safe substance among the general public and increasing cannabis legislation, which has been shown to affect the development of psychosis among cannabis users(54). Thus, investigating changes in associations between psychotic disorders and cannabis us-related variables over time is warranted. The current study shows that the prevalence of self-reported psychotic disorders increased among US adults between 2001-2002 and 2012-2013. Further, results demonstrate that all non-medical cannabis use-related variables were associated with self-reported psychotic disorders in 2012-2013. However, only non-medical cannabis use and CD were associated with self-reported psychotic disorders in 2001-2002, with CD showing a significant strengthening in its association with self-reported psychotic disorders across these time periods. Findings also indicate that residing in a state with MCL had no effect on study results.

Our finding that the prevalence of past-year self-reported psychotic disorders significantly increased between 2001-2002 and 2012-2013 adds to the literature in that it is the first reported change in prevalence of self-reported psychotic disorders drawing on large-scale, nationally representative samples of US adults. Previous studies reporting changes in prevalence of psychosis were limited in that they were derived from older non-US data, mainly based on administrative hospital records, primarily from inpatient admissions(10, 13-17). These studies may have underestimated the true prevalence of psychosis, considering the improved outpatient therapeutic modalities that may have lowered admission rates for psychotic individuals over time(10). Considering that the NESARC and NESARC-III survey items about psychosis did not differentiate between types of psychotic disorders, we could not account for time trends of specific disorders, nor was it possible to differentiate between prevalences of primary and secondary psychotic disorders. Future studies that would provide a more accurate portrayal of the change in prevalence of psychosis in the US should account for specific psychotic disorders, utilize data from both inpatient and outpatient settings, and employ measures of psychosis that are not self-reported.

Our finding that self-reported psychotic disorders were significantly more prevalent among any past-year cannabis users compared to non-users in both surveys is aligned with results from numerous past studies(23, 24, 55, 56) and adds to the existing literature by reporting prevalences of psychotic disorders among past-year adult cannabis users. These findings inform clinicians and policy makers of the increased likelihood of psychosis among these individuals. It should be noted that self-reported psychotic disorders were significantly associated with frequent cannabis use only in the second survey included in this study; however, this association was not significant when non-users were excluded from the model. This differed from previous findings indicating a dose dependent association between cannabis use and psychotic disorders(27) and should be further investigated. While not possible with the available data, a study design that would allow assessment of a true dose-response, such as a linear regression model incorporating cannabis use frequency and quantity as continuous variables, would shed further light on the matter.

Study findings indicate that cannabis-dependent users are at a substantially increased risk of reporting being diagnosed with a psychotic disorder compared to non-dependent cannabis users, a finding that is also reflected in previous non-US studies(31, 32). Notably, the highest absolute prevalence of self-reported psychotic disorders in this study, nearing 11%, was seen in past-year cannabis users describing DSM-IV CD in the 2012-2013 survey. Findings from sensitivity analyses show that both cannabis withdrawal and CUD with withdrawal (a combination that is closer to the DSM-5 diagnostic criteria for CUD) were also associated with self-reported psychotic disorders in both surveys. One plausible explanation for the higher rates of self-reported psychotic disorders among those with cannabis dependence in 2012-2013 compared to 2001-2002 is the increase in availability of high-potency cannabis products, which have been associated with higher prevalence of psychosis(27, 28). Furthermore, in recent years, more cannabis products that are mixed with other psychosis-inducing substances (e.g., PCP) have appeared, further increasing the risk of psychosis among those with cannabis dependence. Further studies should investigate these possibilities.

Study limitations are noted. First, self-reported psychotic disorders were indicated by a single item rather than physician assessment, as in a previous NESARC study (33). While future national studies of substance use should measure psychotic disorders more extensively, a growing number of studies have explored the validity and reliability of various self-reported measures of psychotic disorders, including the current study’s measure, and reported prevalences that are similar to studies using clinical diagnoses (46, 57, 58). Furthermore, compared to other large-scale national surveys, such as the National Survey on Drug Use and Health (NSDUH), which included self-reported psychosis in a broad measure of “severe mental illness”, NESARC is the only national epidemiologic study that utilized a psychosis variable, separately. Second, cannabis use variables could be subject to social desirability bias, since they were based on self-report(40). Third, although much evidence supports the direction of effect modeled, directionality cannot be determined in cross-sectional data. Additionally, since DSM-IV mental disorders were diagnosed in NESARC, while DSM-5 diagnoses were made in NESARC-III, we could not adjust for the presence of other psychiatric disorders. If national data with consistent DSM or ICD mental disorder diagnoses over time can be found, studies should explore such adjustments. Finally, as household surveys, the NESARC and NESARC-III did not include medically institutionalized participants (perhaps less likely than the general population to use cannabis), or incarcerated participants (perhaps more likely to use cannabis). Thus, study results are not generalizable to these populations.

## Conclusions

The prevalence of self-reported psychotic disorders in the adult US population has significantly increased from 2001-2002 to 2012-2013. Several cannabis use-related variables were significantly associated with self-reported psychotic disorders, specifically in more recent times, with little evidence that findings were driven by MCL effects. Cannabis dependence and cannabis use disorders may be helpful factors in identifying individuals who are more likely to suffer from psychotic disorders. This information can inform clinicians and addiction specialists as to the need in appropriate early interventions and therapeutic modalities for these individuals. Although not directly examined in the current study, policy makers should be aware of the increase in cannabis use and CUD among US adults, due to MCL and RCL, and the subsequent rise in cannabis-related outcomes, such as psychotic disorders.

## Data Availability

NESARC-III Datasets are available for investigators to request access through the National Institute of Alcohol Abuse and Alcoholism.

## Acknowledgments

Funding is acknowledged from the National Institute on Drug Abuse (R01DA034244, Hasin) and the New York State Psychiatric Institute.

